# Assessing the impact of Covid-19 on Nurturing Care in Nairobi Slums: Findings from 5 rounds of cross-sectional telephone surveys

**DOI:** 10.1101/2024.05.08.24307078

**Authors:** Robert C Hughes, Silas Onyango, Nelson Langat, Ruth Muendo, Rachel Juel, Elizabeth Kimani-Murage, Zelee Hill, Betty Kirkwood, Sunil S Bhopal, Patricia Kitsao-Wekulo

**Affiliations:** Department of Population Health, Faculty of Epidemiology & Population Health, London School of Hygiene & Tropical Medicine, Keppel Street, London WC1E 7HT; Maternal and Child Wellbeing Unit, African Population and Health Research Center, Nairobi, Kenya; University College London, Epidemiology and Public Health, Institute of Global Health, London, UK; Newcastle University, Population Health Sciences Institute, Faculty of Medical Sciences, Newcastle upon Tyne, Tyne and Wear, UK UCL

**Author notes:** These authors share last authorship (Betty Kirkwood, Sunil S Bhopal, Patricia Kitsao-Wekulo).

**Keywords:** Early Childhood Development, Urban health, Child health, Childcare, Nurturing Care, Covid-19

## Abstract

**Introduction:** This study investigates the multifaceted impacts of the Covid-19 pandemic on early childhood in three of Nairobi’s informal settlements or slums. Focusing on the first five years of life, a critical period for human capital development, we analyse how Nurturing Care across five domains (Health, Nutrition, Responsive Caring, Early Learning, Security and Safety) was influenced by the pandemic and the mitigation measures that were implemented.

**Methods:** Using a longitudinal design, we conducted five rounds of cross-sectional surveys (with between 578 and 774 respondents in each) over 13 months, correlating with different phases of the pandemic and varying levels of Covid-19 restrictions.

**Results:** Our findings reveal significant disruptions in healthcare services, particularly pronounced in early rounds with missed vaccinations (reported by 1 in 5 parents of infants) and therapeutic healthcare seeking (missed by up to 21% of families). The study also highlights persistent food and nutrition insecurity, with a large majority of families struggling to feed their children (up to 72% in Round 1) due to financial constraints. Economic shocks were near-universal, with widespread losses in income and employment reported; 99.7% of respondents reporting earning less since the start of the pandemic. The use of paid childcare initially plummeted but showed a resurgence over time (up to 21% usage by Round 5) as the pandemic and control measures evolved. Young children were commonly left alone in all rounds, but especially earlier in the pandemic; 24% in Round 1, and at least 13% in all rounds, underscoring the enduring challenges in providing consistent nurturing care in these settings. Very few (less than 2%) of study participants reported direct experience of Covid-19 in their family in any round.

**Conclusion:** Despite adaptations over time and the decrease in reported disruptions, the prolonged economic shock was associated with multiple adverse effects Nurturing Care. The study’s longitudinal scope provides insights into the dynamic nature of ensuring young children in slums thrive during crises, highlighting the need for tailored interventions and policies that address the compounded vulnerabilities of young children in these communities.

## 1. Introduction

Early childhood development (ECD), by which we refer to the first five years of a child’s life, is central to setting a child’s developmental trajectory. In recognition that this period is a critical one for accumulation or loss of human capital the early years have received increasing attention from policy makers (1), funders (2,3) and academics(4) in recent years. This culminated in the launch of the Nurturing Care Framework by the WHO/UNICEF and World Bank in 2018(1).

However, there is limited published literature on ECD in urban areas, particularly in slums or informal settlements^1^, despite the rapid growth of these settings. In particular, little is known about either the provider of care, or what care is provided to young children(5). In addition, the SARS-Cov-2 pandemic from 2020 led to radical disruption of early childhood in almost every country around the world due to both the direct and the indirect effects of the virus and efforts to control it(6). While there has been considerable emphasis and research into the effects of the pandemic on health systems and services (7), there has been little work on early childhood specifically, especially in low- and middle-income countries where resilience to shocks may be most limited(8).

We therefore aimed to track over time how the care of young children in three slums in Nairobi was affected by the pandemic and efforts to control it. We aimed to estimate impacts across all five components of Nurturing Care (Health, Nutrition, Responsive Caring, Early Learning and Security and Safety) alongside cross-cutting impacts(for example household economic impacts) during a year of the pandemic. This study is nested within the broader Nairobi Early Childhood Care in Slums (NECS) study, a detailed mixed-methods exploration of the care of children in Nairobi slums(9).

Our key pre-study hypotheses were: firstly, that the evolving Covid-19 pandemic would be likely to impact on Nurturing Care in slums, and that these impacts would be felt across multiple domains. Secondly, we hypothesised that these impacts may evolve over time, in relation to both Covid-19 case incidence and the stringency of epidemic control measures in force.

## 2. Methods

### 2.1. Setting

The setting for the study was three slums in Nairobi, Kenya: Kibera; Mukuru-Viwandani; and Kawangware. These slums are characterized by high levels of poverty, poor sanitation, inadequate shelter, poor infrastructure, high levels of insecurity, and low rates of formal employment(10,11). These slums were selected for several reasons: firstly, these characteristics meant that they represented a particularly vulnerable group; secondly, because of access to a pre-existing database of residents who had consented to being contacted for surveys (described further in section 2.6); and thirdly, to overlap with the wider Nairobi Early Childcare in Slums Study (NECS)(9).

### 2.2 Study design

Five rounds of cross-sectional surveys were undertaken through computer-assisted telephone interviews (CATI). Participants were parents of children aged under five years selected from a pre-existing database of around 48,000 low-income household contacts living in three Nairobi slums (Kibera (c. 25,000 contacts), Kawangware (c. 13,000 contacts) and Mukuru-Viwandani (c. 10,000 contacts). These were people who had previously participated in surveys/field experiments conducted by the non-profit research firm BUSARA(12) and who had consented to being invited to participate in future studies. These potential participants were initially recruited door-to-door by field assistants.

The first round of data collection was from 10 to 29 November 2020, which corresponded with the peak of Kenya’s second wave of recorded Covid-19 infections. Rounds 2-5 were all in 2021 (Round 2: data 9-29 March; Round 3: 6-25 June; Round 4: 6 September -1 October; Round 5: 29 November-13 December).

Table 1 summarises the prevailing Covid-19 epidemiology and controls in place at the time of each round of data collection.

**Table 1.**
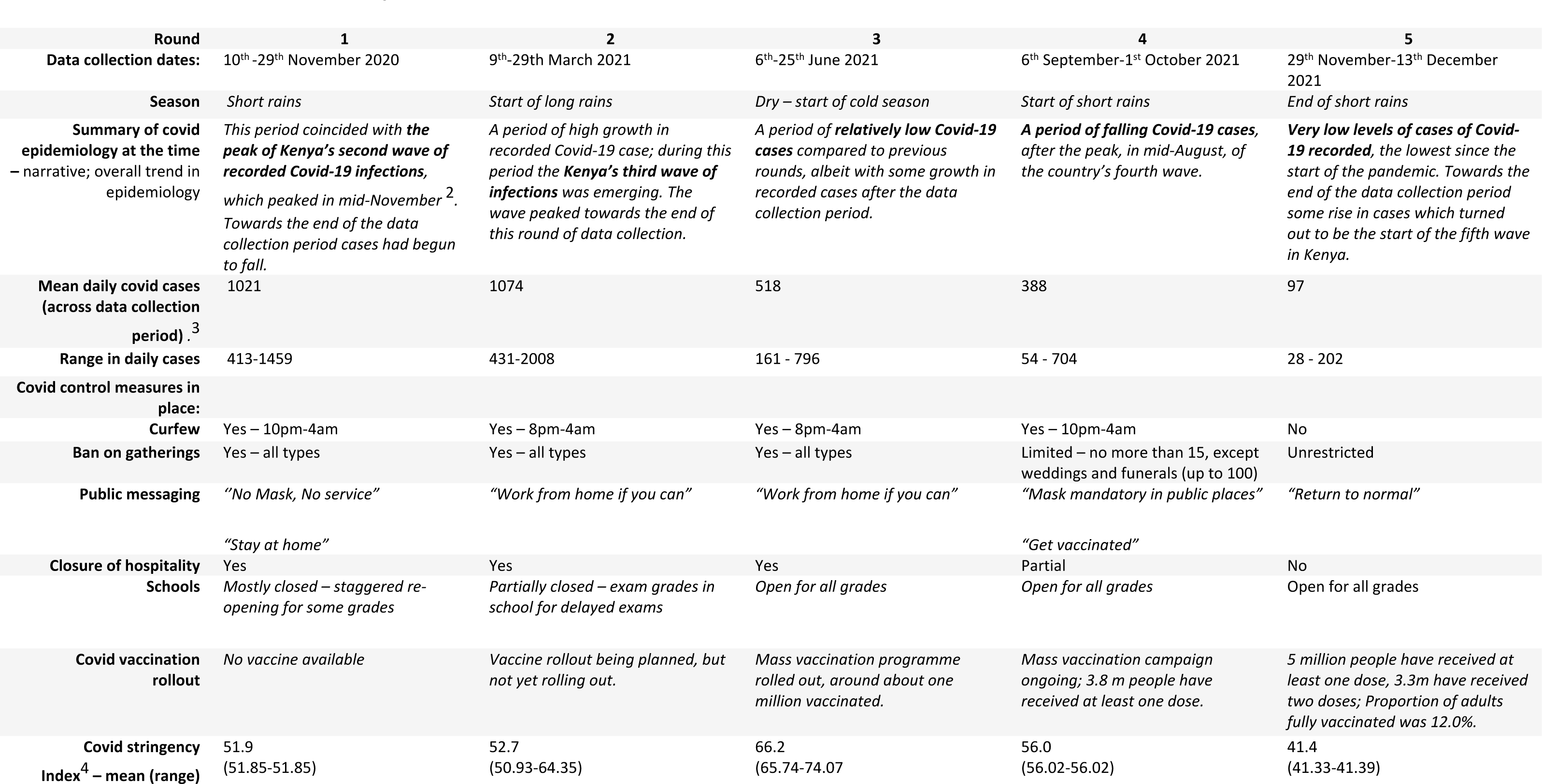
The context for each survey round.

### 2.3 Sampling and recruitment

Survey respondents were selected randomly from the BUSARA database. The recruitment procedure was as follows:

1. A randomly ordered list of potentially eligible respondents was identified based on their being resident in the three slum areas (52% of these were from Kibera, 27% Kawangware, and 21% from Mukuru-Viwandani);
2. An SMS message was sent briefly introducing the research and informing them that they would be called.
3. An eligibility check telephone call was completed, during which, if eligible and interested, the background and rationale for the study were explained, followed by a clear and explicit consent process. This was followed by either immediate completion of survey, or a further scheduled callback at convenient time

Up to five call attempts were made to reach each potential participant each round, at a variety of times during the day. When surveys were interrupted by a dropped call or other interruption, up to five attempts were made to complete the survey over the following days.

Steps 2-3 were repeated in each subsequent round of data collection. If a round was missed, respondents remained in the study, and the same procedures applied in each subsequent round.

### 2.4 Respondent eligibility criteria

Respondents were eligible to participate in the study if they: (i) were 18 years or older; (ii) had a child aged under 5 years old living in their households (or who lived there at some point in the year 2020; in order to avoid excluding children who had moved out of the city due to the pandemic); (iii) were resident in the study setting. In addition, individuals needed to have provided consent to BUSARA within the last 5 years to be contacted with invitations for future research studies.

### 2.5 Data collection

Data were collected by enumerators working for the non-profit research firm BUSARA(12) who had expertise in public health and epidemiology research. The SurveyCTO(13) platform was used for digital data collection which was used to guide call-center operators, (who were experienced in survey work), through the survey questions in Kiswahili. Enumerators had been trained for a total of 12 hours before the start of data collection. They worked from home (due to Covid-19 control measures) with daily check-in video call meetings with supervisors. Data quality was managed through enumerator training and pre-testing, automated range and consistency checks, manual checks on a sample of the data, debriefings, and refresher training. Survey interviews were digitally audio recorded for quality checks (conducted by supervisors on 10% of total surveys in each round).

### 2.6 Survey content

The survey consisted of a set of mostly closed questions. The survey questions were derived from a previously developed conceptual framework and draft household survey tool for the Nairobi Early Childcare in Slums Study(9), supplemented with additional questions aiming to identify the direct and indirect impacts of the Covid-19 pandemic on young children and their families.

Questions covered the following domains:

- Respondent characteristics and survey eligibility
- Child characteristics, including age, and reported sensory or mobility difficulties
- Who was looking after their youngest child, for even a few minutes and for >1 hour in the previous three days.
- Additional questions about the usage frequency and costs of paid childcare amongst those reporting using it
- The Family Care Indicators (FCI) questions about daily activities(14)
- Disruptions to healthcare services or care seeking
- Food and nutrition security, including impacts on breastfeeding
- Perceptions of community safety and security and reports of violence against children
- Levels of concern about the pandemic
- Economic impacts of the pandemic, including loss of work or income
- Help received
- Household assets (only the first time respondents took part)

Where there was more than one child in the house, respondents were asked to focus on the youngest child in the household.

**Error! Reference source not found.** includes the full survey instruments for Round 1 and Rounds 2-5, noting the minor changes that were added following review of Round 1 data quality and free text ‘other’ answers.

### 2.7 Sample size

In Round 1 we planned to recruit an initial 600 participants, allowing for a 20% loss to follow up, with the aim of achieving 480 participants who were followed throughout the study. This was calculated in order to yield estimates of the type of childcare used with precisions of at most +/- 5% including a 25% adjustment for clustering. In practice, we oversampled (by 174, to a total of 774 responses) in Round 1, to allow for higher levels of loss to follow up. In Rounds 2-5, new respondents were recruited following the same procedures as in Round 1.

### 2.8 Data Analysis

Data were analysed using STATA 18(15) and Microsoft Excel365(16). Descriptive summary statistics were calculated to describe the samples for each round and changes between rounds. The frequency of reported impacts across all domains of Nurturing Care and cross-cutting impacts was reported with some indicators (breastfeeding problems, the use of childcare, FCI indicators and children left alone) disaggregated by age of child. Only fully completed surveys were included in the final dataset. A principal component analysis (PCA) was conducted based on a list of 21 reported household assets to generate socio-economic status (SES) quintiles for each unique respondent. Only surveys that were completed fully were shared by BUSARA; i.e. there were no missing data in the dataset we received.

Additional information on Covid-19 epidemiology (drawn from WHO Data Dashboards(17)) and Covid-19 control measures (taken from both the Kenyan Ministry of Health Website(18) and the Oxford Covid Policy Tracker(19)) was collated for use in analyses and interpretation of the survey data.

### 2.9 Public involvement

During the design and inception phase for the wider NECS Study (in February 2020), public engagement meetings were held with community-based organisations in Mukuru-Viwandani. These meetings provided an opportunity to discuss the research methods and questions with the local community, including the planned household survey that informed these telephone surveys. Pre-study visits to the study site allowed for initial discussions about the research questions with parents, childcare providers, and other community members. During preparation of this manuscript emerging draft findings were shared in a community meeting in Nairobi in March 2022, where these findings were discussed with community members, with a focus on implications of the research for local planning and policy making.

### 2.10 Ethics

Informed verbal consent was obtained from all respondents and was recorded for audit purposes. The consent process included provision of information about the purpose of the study, confidentiality, how data would be used and shared, and the right to withdraw consent at any time. Participants who completed a survey round received KES150 (equivalent to US$1.5) for each round of survey completed to compensate them for their costs of participation). The LSHTM Research Ethics Committee (LSHTM Ref: 22692) and Amref Health Africa’s Ethics and Scientific Review Committees (ESRC) in Kenya (Ref: P777/2020) reviewed and approved the study protocol. The National Commission for Science, Technology and Innovation (NACOSTI) provided research clearance. The consent script and anticipated ‘frequently asked questions’ are included in **Error! Reference source not found.**

### 2.11 Role of the funders

The NECS Study was funded by The British Academy (Grant ECE190134) and Echidna Giving who supported a linked Clinical Research Fellowship for RCH. Echidna Giving also provided a ‘Covid Response’ grant for additional costs associated with the Covid-impacts tracker study that is reported on in this manuscript. The funders had no role in the study design, data collection, data analysis, data interpretation, or writing of the paper. The corresponding author had full access to all the data in the study and had final responsibility for the decision to submit for publication.

## 3 Results

### 3.1 Survey and respondent characteristics by round

A total of 1077 participants completed at least one round (774 respondents in R1, 664 in R2, 594 in R3, 528 in R4 and 642 in R5). The flow between rounds is illustrated in **Error! Reference source not found.**, which shows that 774 entered the study at R1; 122 at R2, 58 at R3, 62 at R4, 61 at R5, and that most loss to follow up occurred between R1 and R2. 279 respondents completed all 5 rounds (197 completed 4 rounds; 131 three rounds; 134 two rounds; 336 one round) (Supplementary Table 2).

The median duration of interviews inR1 was 27 minutes, which increased to 34 minutes in R2 (when additional questions were added), and then reduced to 23, 20 and then 17 mins in subsequent rounds (as the number of new recruits fell and familiarity with the questions increased^5^). Most respondents were interviewed on the first call attempt (86-95% in R2-5; data on the number of call attempts was not retained by BUSARA in R1) (Supplementary Table 1).

Table 2 summarises the respondent characteristics by round. Respondents were mostly mothers (52%), fathers (35%) or grandmothers (8%) (see Supplementary Table 3). The mean age of survey respondents was similar across rounds (Range across rounds: 32.0 to 33.8 years old), as was the percent educated to 18 years old or more (Range: 28.3% to 33.4%). Similarly, the proportion of respondents in the lowest socio-economic quintile was consistent across rounds (Range: 20.5% to 17.4%).

**Table 2.**
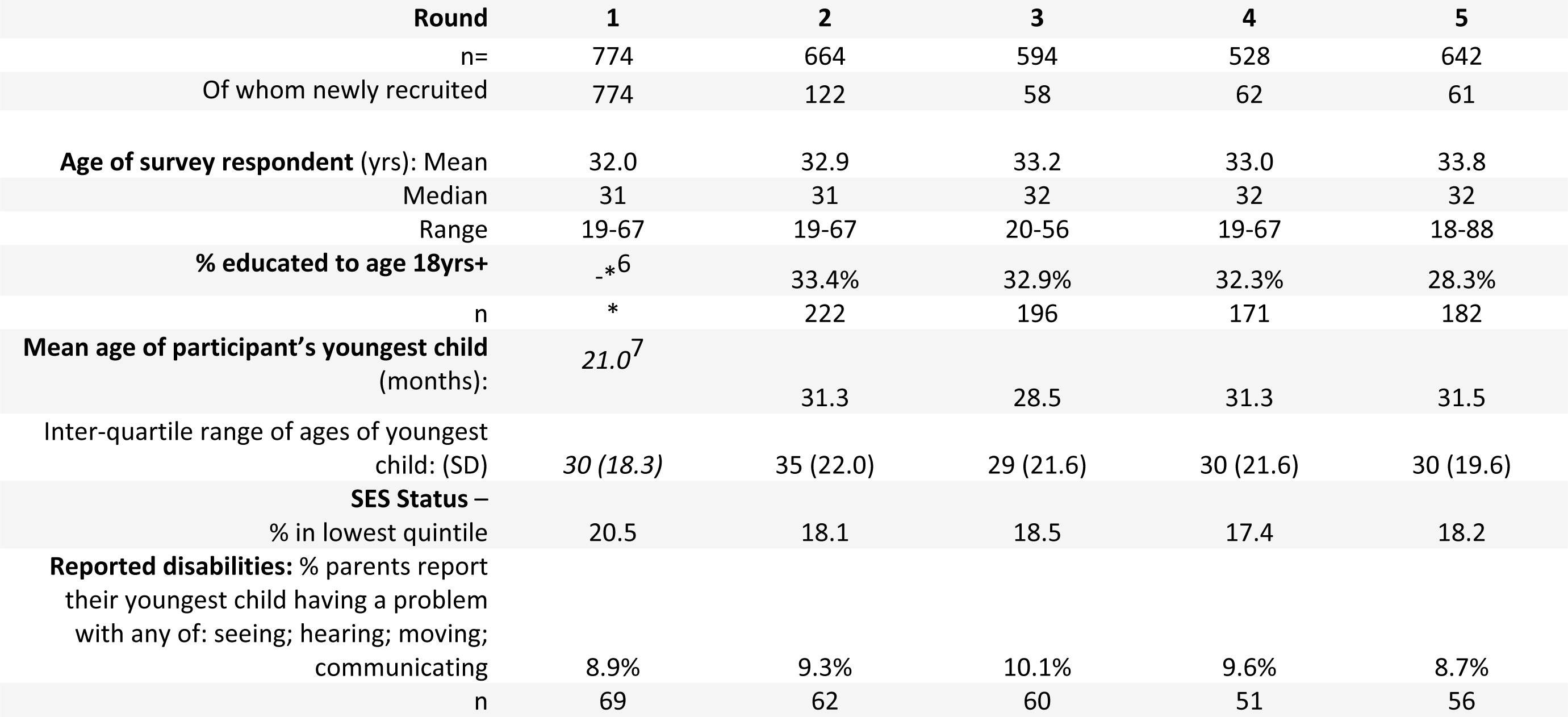
Survey Respondent Characteristics by Round.

In each round 8.7 to 10.1% of respondents reported that the youngest child had a problem with at least one of seeing, hearing, moving or communicating. Problems seeing were most common, followed by communicating and moving (see Supplementary Table 4).

### 3.2 Impacts by Nurturing Care Framework Domain

Table 3A summarises the reported impacts in each round across the domains of Nurturing Care.

**Table 3.**
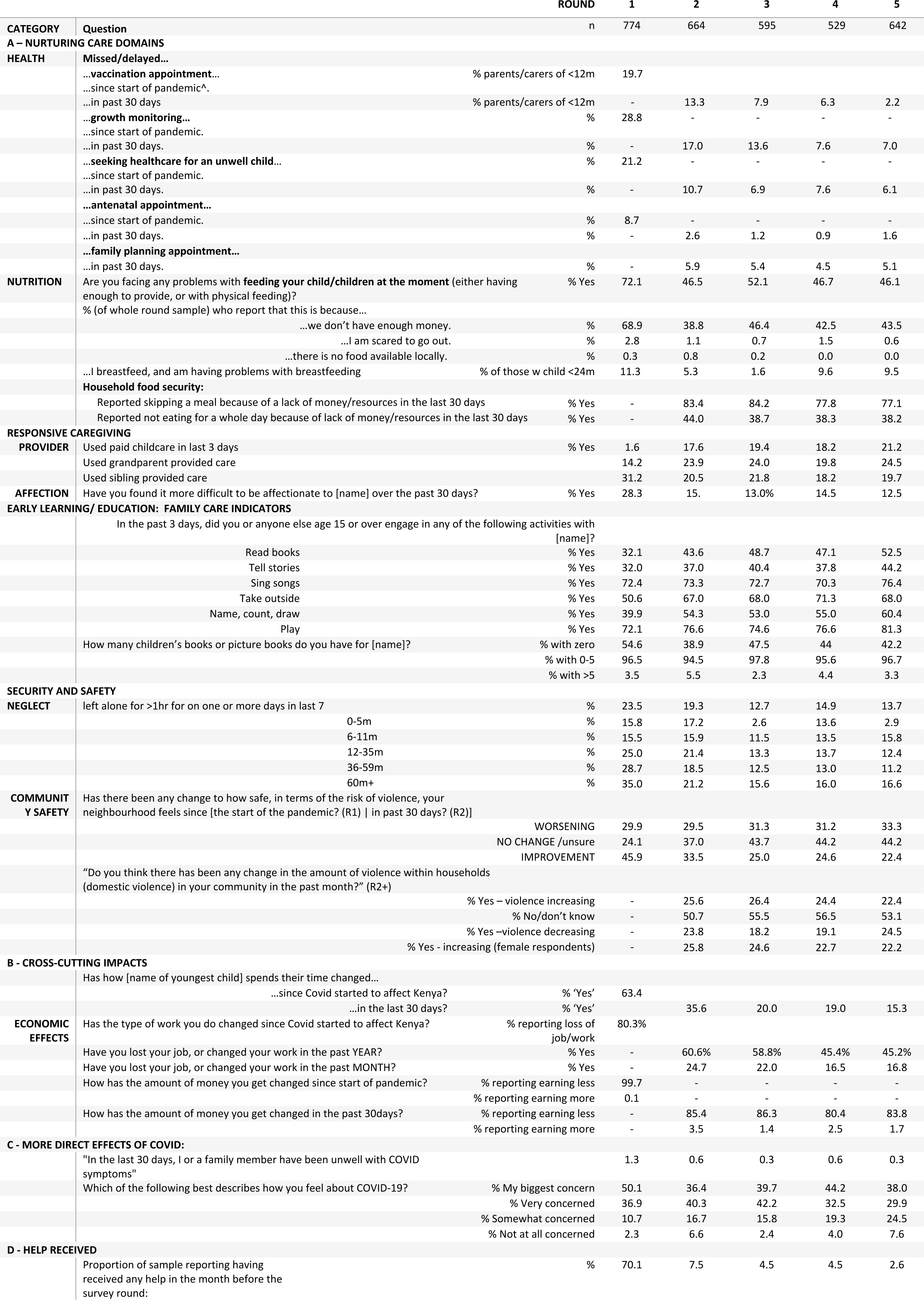
Key Results by Category and Survey Round.

#### 3.2.1 Health

Significant disruptions were reported to a broad set of health services in early survey rounds, and although disruptions continued, there was a reduction in the reported incidence over time. In R1, when asked about disruptions to health services since the start of the pandemic in Kenya, the most frequently reported missed or delayed services was child growth monitoring (reported to have been missed/delayed by 28.8% of respondents) followed by missed vaccination appointments (which was reported for 19.7% of children aged under 12 months old) and seeking care for an unwell child (21.2% reported missing this, across all ages of child). In R1 8.7% of parents/carers reported a missed or delayed antenatal appointment since the start of the pandemic in R1.

In subsequent rounds, respondents were asked about disruptions in the previous 30 days. Across these rounds, there is a largely consistent pattern of declining levels of reported disruption round to round. By R5, 7.0% reported having missed/delayed a growth monitoring appointment, 6.1% delaying/avoiding seeking care for an unwell child, and 5.1% having missed a family planning appointment, and 1.6% an antenatal appointment. Only 2.3% of respondents with a youngest child aged under 12 months reported having missed a vaccination appointment in the last 30 days by R5.

#### 3.2.2 Nutrition

A high proportion of respondents reported problems feeding their child/children in all rounds (R1: 72.1%, R2-5 46.1-52.1%). A large majority of these reported that this was because of having insufficient money (68.9% of respondents reported this in R1), with a much smaller proportion citing being afraid to go out (2.8%) as the reason for their difficulties. Few if any respondents reported food being unavailable locally in any round (0%-0.8%). Amongst respondents with children aged under 24 months, breastfeeding problems were cited by 11.3% in R1, and between 1.6% and 9.6% in subsequent rounds, with no clear trend in over time.

In R2-5 questions from the Food Insecurity Experience Scale (FIES)(20) were included in the survey. In R2, 83% of respondents reported having skipped a meal and 44% reported having not eaten for a whole day due to a lack of money/resources. Reported indicators of food insecurity remained high but dropped slightly in subsequent rounds. By R5, 77% reported having skipped a meal and 38% reported having not eaten for a whole day.

#### 3.2.3 Responsive Care

In R1, only 1.6% of respondents reported having used paid childcare in the previous 3 days. In subsequent rounds the levels of use of paid childcare increased considerably, for example to 17.6% in R2 and to 21.2% in R5. Box 1 summarizes the frequency of use by age, hours and fees paid for childcare reported. Grandparent provided care was common across all rounds, but was lowest in R1 (14.2%), and highest in R5 (24.5%). Sibling provided care was common in R1 (28.3% reported using it) but declined to 12.5% use by R5.

When asked “Have you found it more difficult to be affectionate to [name] over the past 30 days?” 28.3% said yes in R1, 15.6% in R2 and 12.5% by R5.

###### Box 1: Use of Paid Childcare

Figure 2 illustrates the probability of children of different ages using paid childcare in each round. Older children may have more commonly used childcare in early rounds. However, by R4-R5, other than uncommon childcare use amongst children >60m, the probability of children using childcare was similar across all other age groups.

Those who reported using paid childcare were additionally asked about how much it cost, which days it was used, and what hours it was reported to be used for. Most (67-87%) of childcare users in all rounds reported attending for 5 or more days each week (Supplementary table 5). The most common times for using paid childcare were mornings (77-88% of users each round) and afternoons (72-88%), with evening childcare also being frequently used (18-42% of users), but no respondents reporting using paid childcare overnight (Supplementary table 6). The average spend on childcare was similar across rounds, with the median being KES 50-70 (around 0.5 – 0.7 USD) in all rounds (the mean ranged from 67 KES in R4 to 82 KES in R1)(Supplementary table 7).

**Figure 1:**
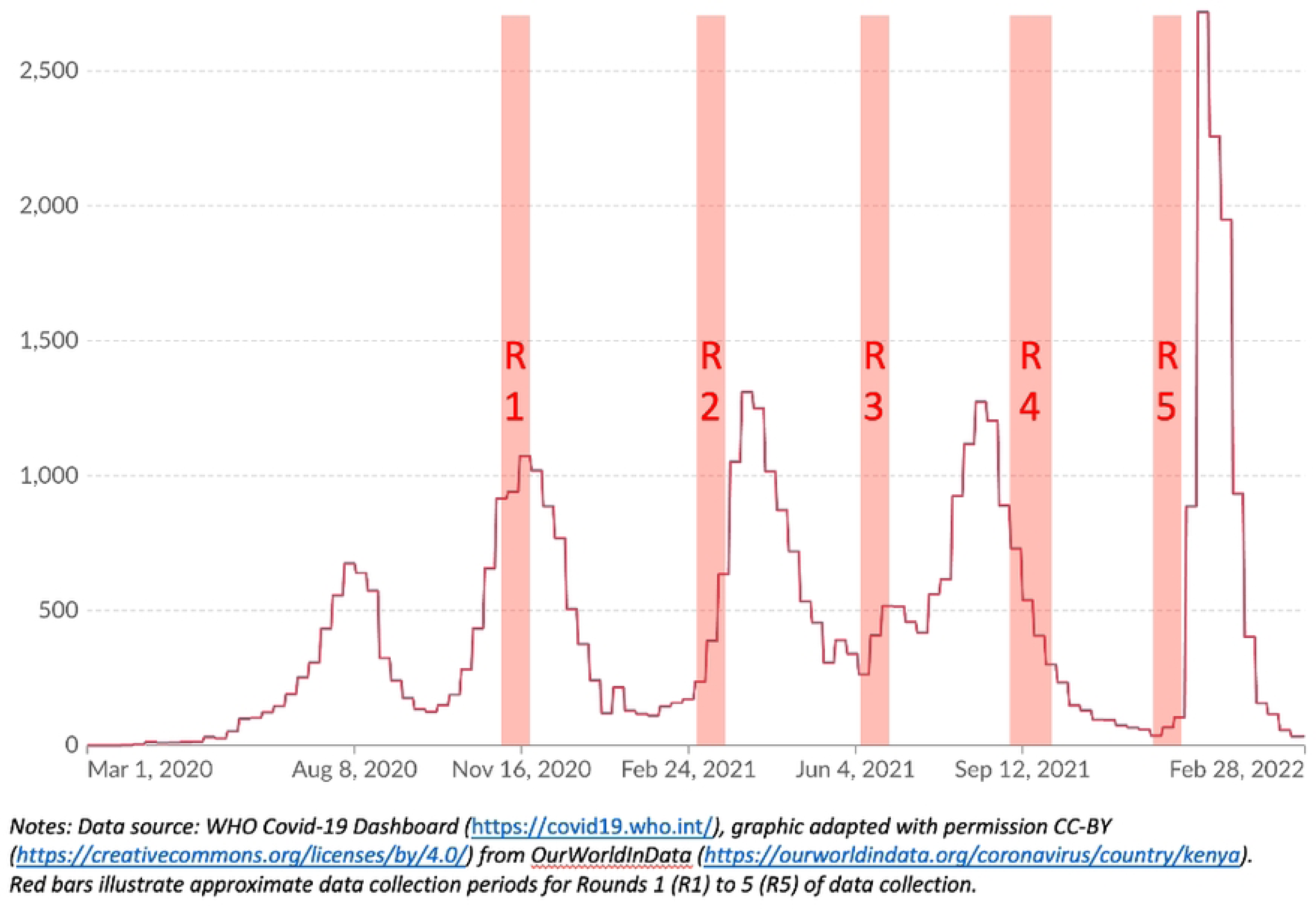
Graph illustrating timing of data collection in relation to 7-day rolling average numbers of confirmed Covid-19 cases in Kenya

**Figure 2:**
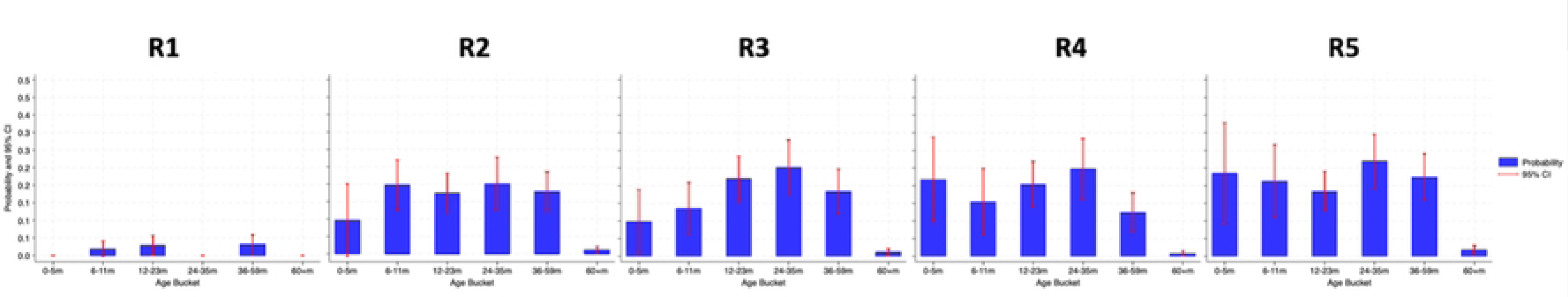
The probability of using paid childcare for each age group in each round

#### 3.2.4 Early learning (and education)

The most commonly reported activities from the Family Care Indicators were playing, singing songs and (especially in subsequent rounds after R1) taking children outside: 72-81% of children were reported to have had a parent or someone over 15 years old playing with them in the previous 3 days; 70-76% reported to have sung songs; and although in R1 only 51% reported having taken their youngest outside in the previous 3 days, in R2-R5 67-71% reported having done so.

Figure 3 is a heatmap illustrating the percentages reporting each Family Care Indicator for each age group for each round. Some activities were reported more frequently with older children (reading, counting/naming/drawing, and telling stories) and singing was more common with younger children. It also appears that there was a modest increase over time in several activities that was not obviously age confounded.

**Figure 3:**
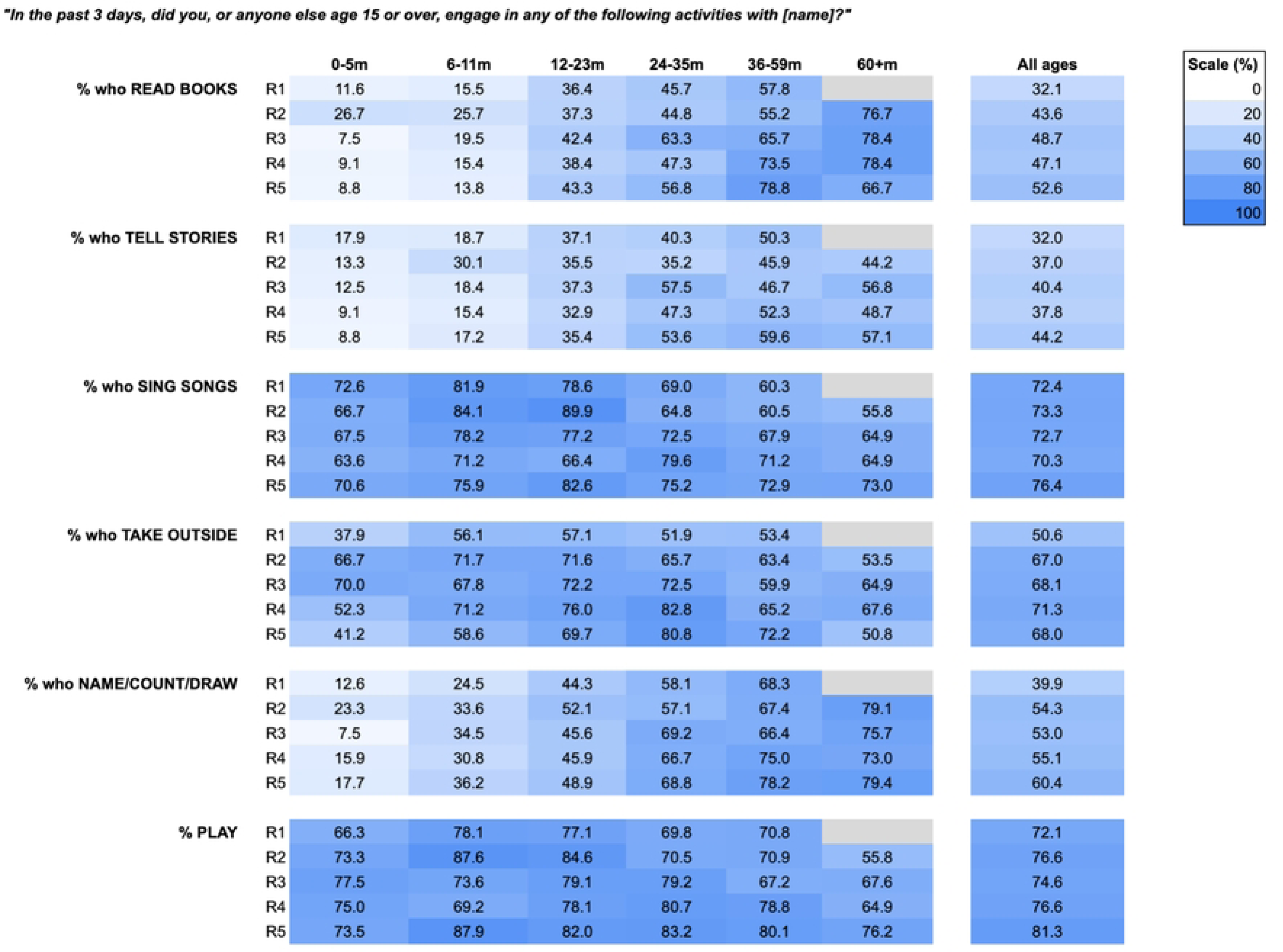
Heatmap illustrating % of respondents reporting ‘yes’ to family care indicators by age

There were few books reported to be in the home; a high proportion of respondents in all rounds reported having no children’s books or picture books (38.9%-54.6%); only 2.3-5.5% reported owning more than 5 books.

#### 3.2.5 Security and Safety

In R1, 23.5% of respondents reported having left their child alone for more than an hour on at least 1 day in the past week. This reduced to 19.3% in R2, and 12.7%-14.9% in R3-5. Although the practice of leaving children alone seemed to be more common with older ages of children, the % of very young children (those aged under 6 months old) reported to be left alone was still high; up to 17% in R2 (Figure 4).

**Figure 4:**
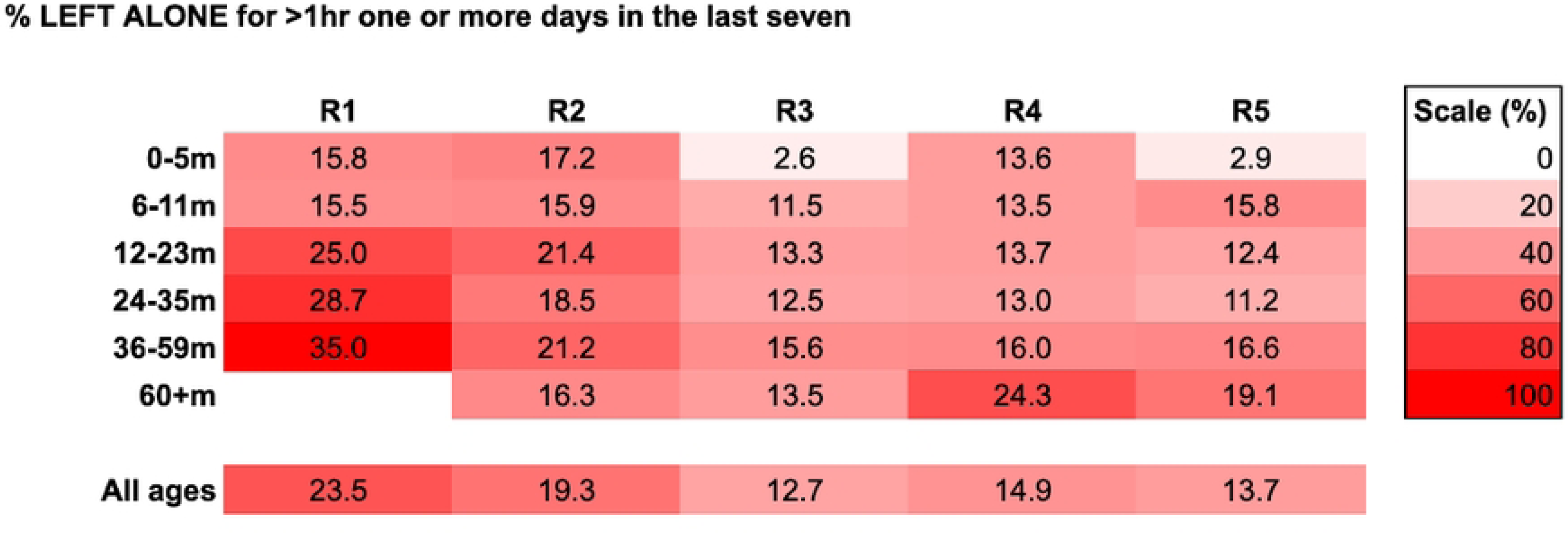
Heatmap of % of children reported to be left alone by age group and round.

Being left under the care of children was also common, with 44.9% reporting in R1 having left their youngest child under the care of someone aged under 15 years old for more than 1 hour in the past week. In R2, 47.1 reported leaving a child with an under 15-year-old, and 33.5% leaving them with an under 10-year-old. The frequency of being left under the care of a child declined from R1 to R3, and then remained relatively consistent, with 37.1% and 26.9% being left alone with an under 15-year-old and an under 10-year-old respectively by R5.

When asked about community safety “Has there been any change to how safe, in terms of the risk of violence, your neighbourhood feels [since the start of the pandemic – R1 / in the past 30 days – R2-5]?” a mixed picture emerged, and this evolved over time. Across all rounds, up to one third of respondents reported a worsening of community safety (range: 29.5%[R2] to 33.3%[R5]). However in earlier rounds a large proportion (45.9% in R1, 33.5% in R2) of respondents suggested safety had improved. By R5, the largest proportion (44.2%) reported that safety had stayed the same or ‘Don’t know’.

Reported violence towards young children was similar across rounds, ranging between 6.3% in R1 to 4.8% in R4 responding ‘Yes’ to the question “In the past month, has anyone been angry or violent towards your youngest child?”. Amongst children under 2 years-old, the rate varied between 1.3% (R2) and 5.4% (R3) across all 5 rounds.

In rounds 2-5, an additional question asked about perceptions of any change in household/domestic violence; in all but R5, a slightly higher proportion of respondents suggested that domestic violence was increasing (25.6%-24.4% R2-4), than suggested it was falling ((23.8% - 19.1% R2-R4). In all rounds around half of respondents replied that it was not changing or ‘Don’t know’. Amongst female respondents (those reporting their relationship to the child was one of being a mother, grandmother or aunt) the rates of increased perceptions of domestic violence were similar to those in the whole sample (25.8%[R2] to 22.2%[R5]).

## 4 Cross-cutting impacts

Table 3B summarises reported cross-cutting impacts, including effects on how young children spent their time and on household income and work. In R1, nearly two thirds (63.4%) reported that the pandemic had led to a change in how their young child spent their time, with the top reported reasons for this being: the closure of childcare; lack of money making childcare unaffordable; the impact of partial reopening of schools; and reductions in contacts with peers/relatives. Over the course of survey rounds, there was a tendency towards lower levels of reported disruption, with only 15.3% reporting a change in the past 30 days by R5, compared to 35.6% reporting this in R2.

Economic impacts of the pandemic were reported to be widespread; in R1 80.3% reported a loss of job/source of income since the start of the pandemic, and 99.7% of respondents reported earning less than in the pre-pandemic period. When asked in subsequent rounds about change in the past 30 days between 80.4% and 86.3% continued to report a reduction in income each round. Very few respondents (0.1-3.5%) reported an increase in earnings in any round.

### 3.3 Reported incidence and concern about Covid-19

When asked about any other impacts of the pandemic, a small proportion of respondents, between 1.3% in R1 and 0.3% in R3, reported that they or a family member had been unwell with Covid-19 symptoms (Table 3C). Levels of concern about Covid-19 were very high initially (with 87% of respondents saying that Covid-19 was their “biggest concern” (50%) or that they were “very concerned” about it (37%)), and although this declined over the course of the data collection period, even by R5, 38% were still saying that Covid-19 was their biggest concern.

### 3.4 Help received

The proportion of people reporting having received any help was significantly higher in R1(70.1%) compared to R2 (7.5%) and later rounds, falling to 2.6% in R5. Most help was reported to have been received in the earlier time periods surveyed, and was in the form of being given information (reported to as received by 41.5% in R1), being given support for their mental wellbeing (for example someone sitting with you, talking with you), (28.8% in R1), and donations of masks (28.6% in R1).

## 4 Discussion

**Principal findings:** There are several notable features of the reported impacts of the pandemic on Nurturing Care in slums. Firstly, the impacts were broad in their extent, affecting all components of Nurturing Care. We show that healthcare was radically disrupted, especially earlier in the pandemic, for example with 1 in 5 families with a child aged under 12 months reporting having missed a childhood vaccination appointment in R1, and a similar proportion (21%) of families of children aged up to five years reporting having missed or delayed seeking healthcare for an unwell child since the start of the pandemic. We also identified high levels of reported food and nutrition insecurity, including 4 in 10 respondents having not eaten for a whole day in all the rounds where this question was asked. High levels of disruption to responsive caregiving were reported, including the use of paid childcare which was reported by less than 2% of respondents in R1. It was common for children, including very young children, to be left alone. This was especially the case earlier in the pandemic (24% of children under 5 years and 16% of those aged under 6 months old were left alone for a least one hour on at least one day in the past week in R1).

Secondly, there was considerable change in reported Nurturing Care over time as the pandemic and control measures evolved from an initial strict ‘lockdown’ with closed schools and workplaces to a near-return to normal by the final round of data collection. The most notable changes over this period were the return to widespread use of paid childcare (which increased to more than 1 in 5 reporting using it by R5). Parents/carers of even very young children used paid childcare regularly (usually for five or more days a week), paying very low fees (commonly around KES50 or 0.5USD/day) to do so. We also found **r**eductions in the frequency of reported disruptions to healthcare over time (by R5 only 2.3% of parents of infants reported having missed a vaccination appointment in the last 30 days) and a decline in the incidence of lost jobs/incomes. However, other impacts were reported to be persistent. For example, even by R5, 14% of children aged under 5 were still being left alone on at least one of the last seven days, and high levels of food insecurity continued to be reported.

Underlying all of these was an almost universal experience of a significant economic shock, with almost all respondents reporting a loss of work or income, especially earlier in the pandemic; 99.7% of respondents reporting having experienced a reduction in work or income in R1. Strikingly, despite high levels of concern about Covid-19, especially in early rounds, very few (less than 2%) of study participants reported direct experience of Covid-19 in their family in any round.

**Strengths** of this study include its longitudinal design; five survey rounds spread over a year provide insights into temporal changes and trends in the nurturing care of young children in these slums. In addition, the explicit focus on people living in slums provides valuable insights into an often-underrepresented demographic in ECD research(21). Finally, the multi-domain approach, encompassing all components of nurturing care, offers a broad assessment of the impacts of the pandemic and attempts to control it on young children in these settings.

**Limitations** include a reliance on self-reporting, which may introduce biases or inaccuracies in the data, including both recall bias and the potential for response bias including strategic misrepresentation (if respondents thought that replying in certain ways might lead to allocation of resources to their predicament)(22). The reliance on a pre-existing database of potential respondents, all of whom had access to a mobile phone, may also have introduced selection bias including an over sampling of better off participants. In addition, the changes in the respondent pool over the study period could affect the consistency of the findings (although efforts were made to account for the risk of age confounding in the analysis of results). Finally, the focus on three slums in Nairobi limits the extrapolation of findings to other settings.

**Situating these findings within the broader literature:** These findings are broadly consistent with other research that has been published on the impacts of the pandemic on the care of young children, albeit with few comparators from informal settings and studies which have tracked impacts over time. For example, health service disruption has been documented through time-series analyses in Kenya which showed that the biggest declines in activity were in outpatient visits and childhood immunisations, with some rebound as the pandemic evolved(23). Several studies have documented the radical economic shocks precipitated by the pandemic, including globally/regionally(24) and within Kenyan slums particularly, where the pandemic and resulting government policies were reported to have had devastating consequences on the livelihood of slum dwellers who were “left to choose between life and livelihood”(25), “violating” their human right to food(26). Such findings were replicated in Bangladesh, where the impacts on household economics (27,28) and the mental health of caregivers (27) were stark. In addition, the quantitative findings presented in this paper are consistent with our qualitative research with parents living in Nairobi slums during the pandemic, which described socio-economic impacts which were deep, protracted, and widely felt (29,30).

One review that looked at the impacts of the pandemic on Nurturing Care found a bias within the published literature towards quantitative studies in high-income countries and those focused on caregiver mental health(8) However, a wide range of UNICEF reviews has documented a broad set of global impacts on children arising from the pandemic, including early learning losses(31), nutrition impacts(32), disruption to child protection(33) and health services(34,35). Few studies have looked at trends over time as the pandemic evolved, but some have sought to estimate how disruptions to health(36) and broader early childhood adversities precipitated by the pandemic may translate into short- and long-term impacts(37).

By the final round of our data collection, around 1 in 5 children were attending paid childcare regularly. This is lower than another estimate in a different Nairobi slum (Korogocho), in which 46% of employed and 23% of unemployed mothers reported using paid childcare for daytime care for their children aged 1-3 years (38). This variance could be related to differences in sampling, the ongoing impacts/legacy of the pandemic, differences between the settings or differences in how the question was framed and asked. The fees charged that we found are similar to another recent Kenyan study, and highlight the limited resources in the childcare system(39).

**Unanswered questions and future research:** to build on this research it would be valuable to examine how the disruptions noted in this study translate into impacts on child outcomes. In the short-term, this could include child health and development outcomes and then school readiness, but longer term it will be important to understand how the early childhood adversities experienced by this cohort of children, and parents, translates into later life learning, earning and wellbeing. That said, in many ways it is more important and urgent that further implementation research is conducted to develop and test intervention strategies, potentially building on the potential of the childcare platform, to mitigate from some of these disruptions. In addition, these findings underscore the need for better research to characterise the childcare system, including the economics, policies and perspectives of key stakeholders involved, in and beyond Kenya(40,41).

**Policy implications:** This research suggests a number of ways in which policy and practice can better serve young children growing up in slums or informal settlements. Firstly, as noted by others(26) people, and especially children, growing up in slums deserve a much greater consideration in public including pandemic policy making. The application of blanket policies to very different settings ought to be avoided(25), and in particular the practicalities and impacts of attempting to ‘lockdown’ a slum – where most residents shop daily for necessities, and many rely on daily wages – in particular ought to be better recognised(42). Secondly, the data presented here and in related studies yimplies an urgent need for implementation of ambitious and well-resourced interventions to address both the underlying vulnerabilities experienced by young children growing up in slums, and the ways that there were amplified by the pandemic and attempts to control it. Paid childcare – a platform that seems to be widely used yet significantly under-resourced - may provide an intervention opportunity to address some of the risks identified in this research(43). Finally, noting that many of the risks to the wellbeing of children we predicted early in the pandemic with accompanying calls for mitigation of harms to young children(37), given that these calls seem to have rarely been heeded, it may be that more research and innovation is needed into the structural changes needed to better bring the next generation’s voices and rights into decision making(44).

## 5 Conclusion

The early years are a period of both opportunity and vulnerability. This study explored how the care of young children growing up in slums was affected by the Covid-19 pandemic and the control measures brought in to control it. A deep and broad set of impacts across all domains of Nurturing Care were evident. While some of the disruptions appeared to resolve over the five rounds of data collection, others – most notably those related to the economic shocks related to the pandemic – persisted. Provision of paid childcare was effectively suspended in the early part of the pandemic, but usage recovered over time. Collectively, these findings imply an urgent need for greater policy attention being paid to the care of young children growing up in slums, especially at times of crisis.

## 6 Conflict of Interest

The authors declare that the research was conducted in the absence of any commercial or financial relationships that could be construed as a potential conflict of interest.

## 7 Author Contributions

RH, SB, BK, PK-W, and ZH were responsible for conceptualizing the study. RH played a key role in planning the paper, leading the analysis, and writing the first draft of the paper. RM, with PK-W, SO, ZH and SB supervised and supported the data collection. RH and RJ completed data cleaning and initial quality checks. RH and SB and then all authors reviewed draft manuscripts. All authors edited the draft and contributed important intellectual content. The final manuscript was approved by all authors.

## 8 Funding

The funding for this work was provided by the British Academy (Grant number ECE190134) and Echidna Giving, which supports RH through a linked Clinical Research Fellowship and which provided a Covid Response Grant to support of this study.

## Data Availability

The datasets analysed for this study can be found in the LSHTM Data Compass site for the NECS Study(45).

https://datacompass.lshtm.ac.uk/1780

## Acknowledgments

We would like to firstly thank the study participants who completed the telephone surveys reported here at a time of considerable disruption. We would also like to acknowledge and thank the data collection team at BUSARA, in particular Jennifer Adhiambo, and the critical administration teams at both APHRC and LSHTM (Antonio Aparicio). We would also like to thank both Echidna Giving and The British Academy for their vital support for this and wider work on childcare in slums. Finally, we would like to thank Dr APS Munro for assistance in extracting Oxford covid stringency index data.

## Footnotes

1 In this paper we have largely adopted the approach and terminology as used in the *Lancet* series on ‘The health of people who live in slums’, which defines the term ‘slums’ as distinct from ‘informal urban areas’ based on neighbourhood effects, many of which seem relevant to the discussion around childcare(4). Nevertheless, our experience is that the terms ‘slum’ and ‘informal urban areas’ are often used somewhat interchangeably, and at times we have used both terms.

2 https://www.health.go.ke/wp-content/uploads/2020/11/NERC-MOH-CS-COVID-RELEASE-15.11.2020e-2.pdf

3 *Data obtained from the Ministry of Health official website* https://www.health.go.ke/press-releases/

4 See COVID-19 Government Response Tracker | Blavatnik School of Government (ox.ac.uk)

5 Questions about household assets were only asked in the first round that a respondent took part in.

6 This question was asked differently after Round 1, as there appeared to be some confusion in how to responded (Participants were asked how many years of schooling they had completed, but frequently reported the age that they left education)

7 In Round 1, insufficient range checks and age verification questions were in place in the survey instrument; something that was corrected in subsequent rounds. This number is therefore reported, but may need to be interpreted with caution.

## Notes

### Competing Interest Statement

The authors have declared no competing interest.

### Author Declarations

The LSHTM Research Ethics Committee (LSHTM Ref: 22692) and Amref Health Africa’s Ethics and Scientific Review Committees (ESRC) in Kenya (Ref: P777/2020) reviewed and approved the study protocol. The National Commission for Science, Technology and Innovation (NACOSTI) provided research clearance.

